# Association between telomere length and cognitive function among cognitively unimpaired individuals at risk of Alzheimer’s disease

**DOI:** 10.1101/2023.05.29.23290678

**Authors:** Blanca Rodríguez-Fernández, Gonzalo Sánchez-Benavides, Patricia Genius, Carolina Minguillon, Karine Fauria, Immaculata De Vivo, Arcadi Navarro, Jose Luis Molinuevo, Juan D. Gispert, Aleix Sala-Vila, Natalia Vilor-Tejedor, Marta Crous-Bou, the ALFA study

## Abstract

**INTRODUCTION:** Leukocyte telomere length (LTL) is an objective biomarker of biological aging, and it is proposed to play a crucial role in Alzheimer’s disease (AD) risk. We aimed at evaluating the cross-sectional association between LTL and cognitive performance in middle-aged cognitively unimpaired individuals at increased risk of AD.

**METHODS:** A total of 1,520 participants from the ALFA cohort were included. Relative telomere length was measured in leukocytes through qPCR. LTL was residualized against age and sex, and associations with cognitive performance were assessed in accelerated and decelerated biological aging individuals based on residual LTL (rLTL). Interactions with sex and genetic risk of AD were tested.

**RESULTS:** Non-linear associations were found between LTL and episodic memory (EM). Better EM was associated with longer rLTL among women in the accelerated aging group.

**DISCUSSION:** Results suggest a potential role for telomeres in maintaining cognition in aging with sex-specific patterns.

**Highlights:**

● There is a non-linear association between telomere length and cognitive performance.
● Longer telomeres are suggested to have a beneficial effect on episodic memory.
● The association between telomeres and cognitive performance might be sex-specific.

## 1. Introduction

Aging is the primary risk factor for most neurodegenerative diseases, including dementia due to Alzheimer’s disease (AD) ^1, 2^. The number of individuals living with dementia worldwide is expected to increase upon 153 million in 2050, due primarily to population aging ^3^. However, deciphering molecular mechanisms linking aging with neurodegeneration is a complex undertaking, as these two phenomena likely evolve in tandem and mutually influence each other ^4, 5^ .

The human telomere complex consists of tandem repeated short DNA sequences and associated proteins located at the end of chromosomes that protects genomic integrity. Telomere attrition naturally occurs during aging due to cellular division ^6, 7^. However, telomere erosion to a critical length can trigger aging-associated disease by inducing genomic instability, which can lead to cellular apoptosis or senescence ^8^. Thus, telomere length (TL) is considered one of the hallmarks of aging, along with others such as epigenetic alterations or chronic inflammation ^9^. These hallmarks are functionally related to each other and are interconnected with the main determinants of health, collectively contributing to the complex and multifactorial nature of aging ^10^.

In epidemiological studies, TL is commonly measured in leukocytes from peripheral blood (i.e., leukocyte telomere length [LTL]) and it has been proposed as an objective biomarker of biological aging across human tissues ^11^. Shorter LTL has been associated with increased mortality rates and higher incidence of several age-related comorbid diseases associated with AD, such as cardiovascular or metabolic traits ^12–15^. Importantly, while LTL is a highly heritable trait ^16^, recent evidence suggests that it may be modifiable by lifestyle interventions and exposure to environmental factors ^17^.

Shorter LTL has been associated with an increased risk of developing AD ^18,19^ and people living with AD showed shorter LTL than age-matched controls in a meta-analysis study ^20^. However, the underlying biological pathways driving these associations remain to be elucidated. In this regard, LTL has been associated with multiple structural brain endophenotypes of neurodegenerative diseases among cognitively unimpaired (CU) middle-aged individuals from the UK Biobank ^21^. Moreover, longer LTL was associated with a lower risk of incident AD, better cognitive performance, larger hippocampus volume and lower total volume of white matter hyperintensities ^22^.

Nonetheless, the association of LTL with early markers of AD, including cognitive performance, requires further investigation along the *continuum* of the disease. For instance, prior studies have reported a detrimental effect of telomere shortening on cognitive performance in the general population ^23–25^. However, LTL was unrelated to cognitive ability among CU older individuals or those having dementia or incipient dementia ^26^. Besides, LTL at baseline was associated with worse cognitive performance among individuals with high baseline AD pathology at different stages of the disease’s *continuum* from the AD Neuroimaging Initiative ^27^.

Given the conflicting findings in previous studies, the primary objective of this study was to investigate the association between baseline LTL and cognitive performance in a cohort of middle-aged, CU individuals at increased risk of AD. In addition, we explored potential non-linear associations, sex-interactions, and effect modifications related to the genetic risk of AD. The results of our study are expected to shed light into the complex relationship between LTL and AD pathological processes during the earliest stages of the *continuum*.

## 2. Methods

### 2.1. Study participants

The present is a cross-sectional analysis that was conducted in the context of an existing study, the ALFA (Alzheimer and Families) study ^28^. Briefly, the ALFA study (Clinicaltrials.gov Identifier: NCT01835717) includes a total of 2,743 CU participants, aged 45–74, many of them kindred of people living with AD (86% had at least one parent with dementia regardless age at onset, 48% of the participants had at least one parent diagnosed with AD before the age of 75).

Participants were characterized at their baseline visit in 2013–2014 at multiple levels (sociodemographic, anthropometric, clinical, epidemiological, cognitive and neuroimaging). . ALFA exclusion criteria were (1) Cognitive performance falling outside established cutoffs: Mini-Mental State Examination ^29, 30^ < 26, Memory Impairment Screen ^31, 32^ < 6, Time-Orientation subtest of the Barcelona Test II ^33^ < 68, semantic fluency ^34, 35^ (animals) < 12. (2) Clinical Dementia Rating scale ^36^ > 0. (2) Major psychiatric disorders (DSM-IV-TR) or diseases that could affect cognitive abilities. (3) Severe auditory and/or visual, neurodevelopmental and/or psychomotor disorders, significant diseases that could interfere with cognition. (4) Neurological disorders. (5) Brain injury that could interfere with cognition. (6) Family history of AD with suspected autosomal dominant pattern. Further details of ALFA participants can be found in ^28^.

The study was conducted in accordance with the directives of the Spanish Law 14/2007, of 3rd of July, on Biomedical Research (Ley 14/2007 de Investigación Biomédica). All participants accepted the study procedures by signing an informed consent form.

### 2.2. Cognitive performance evaluation

During neuropsychological evaluation at baseline, all participants were administered a cognitive test battery for the detection of early decline in longitudinal follow-up. Episodic memory (EM) was assessed by means of the Spanish version of the Memory Binding Test (MBT) ^37, 38^. In this test, the examinee should learn two sets of 16 written words that share semantic categories by pairs. Four main variables including free and cued recalls in immediate and delayed (after 25–35 min) trials were analyzed: Immediate total paired recall (TPR), immediate total free recall (TFR), delayed total paired recall (TDPR), and delayed total free recall (TDFR). Further cognitive domains were measured using the Wechsler Adult Intelligence Scale (WAIS) IV Coding, Digit Span, Visual Puzzles, Similarities and Matrix Reasoning subtests ^39^. Coding measures, among others, processing speed and attention. The Digit Span subtest evaluates short-term and working memory. Visual Puzzles measures complex visual processing. Matrix Reasoning assesses non-verbal reasoning, and Similarities measures verbal reasoning and abstract thinking. For our study, a modified Preclinical Alzheimer Cognitive Composite (PACC) ^40^ was created by averaging the z-scores of the following variables: MBT-TPR, MBT-TDFR, WAIS-IV Coding, and semantic fluency. Moreover, two additional cognitive composites to assess global EM and executive function (EF) were calculated by creating z-scores for the cognitive measures from MBT and from WAIS-IV subtests, respectively. These global measures were calculated by averaging normalized raw scores of all subtests in each domain ^41^. As with the individual cognitive tests, higher scores in the different composites represent better cognitive performance.

### 2.3. Apolipoprotein E genotyping

Total DNA was obtained from the blood cellular fraction by proteinase K digestion followed by alcohol precipitation. Using the following primers (*APOE-F 5′- TTGAAGGCCTACAAATCGGAACTG-3′ and APOE-R 5′- CCGGCTGCCCATCTCCTCCATCCG-3′*) samples were genotyped for two SNPs, rs429358 and rs7412, determining the possible *APOE* alleles: ε1, rs429358 (C) + rs7412 (T); ε2, rs429358 (T) + rs7412 (T); ε3, rs429358 (T) + rs7412 (C); and ε4, rs429358 (C) + rs7412 (C).

### 2.4. Genome-wide genotyping, imputation, and polygenic risk score

A total of 2,686 participants of the ALFA parent cohort were genotyped, as 57 individuals had to be excluded since blood extraction could not be performed or sufficient DNA could not be obtained to perform the genotyping. After genetic quality control (QC) procedure, a total of 2,527 CU individuals from the ALFA parent cohort were genetically characterized. Imputation of genetic variants was performed using the Michigan Imputation Server with the Haplotype Reference Consortium Panel (HRC r1.1 2016) ^42^ using default parameters and following established guidelines. Polygenic risk scores (PRS) were calculated using PRSice version 2 ^43^. PRSice computes PRSs by summing all SNP alleles carried by participants, weighting them by the SNP allele effect size estimated in a previous genome-wide association studies (GWAS), and normalizing the score by the total number of SNPs included. PRS for AD risk, CSF amyloid-β (Aβ) and PRS-CSFpTau were calculated using summary statistics from recent GWASs. PRS for AD was additionally calculated excluding the *APOE* region (chr19:45,409,011-45,412,650; GRCh37/hg19) (PRS-ADno*APOE*). Results were displayed at a restrictive threshold, 5×10^-6^. DNA extraction, genotyping, genetic QC, and imputation procedures are detailed elsewhere ^44^.

### 2.5. Leukocyte telomere length measurements

A total of 1,660 participants were selected for LTL determinations based on the availability of biological samples (already stored at the biobank) and cognitive assessment (available at in-house databases). Samples were sent to the Harvard Cancer Center Genotyping & Genetics for Population Sciences Facility for LTL determination using a high throughput version of the quantitative real-time polymerase chain reaction (qPCR)-based telomere assay. LTL was measured in a single batch for all samples. LTL was determined by qPCR from the DNA extracted from peripheral blood leukocytes. First, DNA was quantified and normalized, and then the relative LTL was determined by a high-performance version of the real-time qPCR for telomeres. The assay was run on the Applied Biosystems 7900HT Sequence Detection System (Foster City, CA, USA). Laboratory personnel were blinded to participants’ characteristics, and all assays were processed in triplicate by the same technician and under identical conditions. The average relative LTL (i.e., Exp ddCt) was calculated as the exponentiated ratio of telomere repeat copy number to a single gene (36B4) copy number corrected for a reference sample.

Sample triplicates coefficient of variation (CV) ranged between 0.15-14.6. Samples with triplicate-CVs above 15% were removed for subsequent analysis (3 samples). The intra-set CVs ranged between 0.97-1.14. The average CV was 8.36% for the whole assay, which passes the internal standard quality controls. Forty-five samples failed the assay (2.6%), of which 21 were expected to fail due to low concentration of DNA after the DNA quantification. A total of 48 samples did not fail but presented higher cycle threshold (Ct) values than they should be (Ct > 26 for telomere and Ct > 29 for 36B4) (N = 48). Additionally, *APOE*-ε2ε4 individuals (N = 30) were removed from the analyses. Outliers were detected and removed based on the Grubbs test (N = 5), leaving reliable data for a total of 1,532 after quality control.

### 2.6. Statistical analysis

LTL values were natural log-transformed and normalized by computing z-scores. LTL was residualized (i.e., rLTL) against chronological age at LTL measurement and sex using a linear regression model. rLTL was considered a standardized measure of biological aging in the study population: greater rLTL values were indicative of longer telomeres than expected for a given chronological age (irrespective of sex) and *vice versa*.

Initial analyses showed suggestive non-linear relationship between rLTL and cognitive performance in the study sample. In order to capture non-linear effects, categorization of the sample was performed by (1) using natural cubic splines to model the association between rLTL and cognitive performance ^45^ and (2) selecting the knots based on the lowest Bayesian information criteria [BIC] ^46^. The most parsimonious description of the rLTL-cognitive performance association was found for a division based on the percentile 50^th^ (according to the BIC, see **Supplementary Figures 1-3**). Thus, the sample was classified in two groups based on the percentile 50^th^ of the rLTL: accelerated biological aging group (i.e., rLTL < percentile 50^th^) and decelerated biological aging group (i.e., rLTL > percentile 50^th^).

The cross-sectional association between rLTL and cognitive performance was tested in the whole sample and separately in the accelerated and decelerated biological aging groups. Generalized linear models with Poisson/quasi-binomial distributions and linear regression models were implemented to assess the association between rLTL and cognitive subtests as well as rLTL and cognitive composites, respectively. All models were adjusted by age, sex, education and *APOE*-ɛ4 status.

Interactions with genetic risk for AD and sex were tested by including interaction terms for sex, *APOE*-ɛ4 status, PRS-ADno*APOE,* PRS-CSFAβ *and* PRS-CSFpTau in the models. Stratified analyses by sex were conducted. In those analyses, LTL was separately residualized against chronological age in women and men. Sensitivity analyses were conducted by further adjusting linear models by systolic blood pressure (SBP), PRS-ADnoAPOE and parental history of AD.

A false discovery rate (FDR) multiple-comparison correction was applied following the Benjamini-Hochberg procedure ^47^ for all analyses. FDR-*P* value < 0.05 were considered statistically significant. Unadjusted *P* value < 0.05 were considered nominally significant. All analyses were conducted under R software, version 4.2.3 ^48^. Descriptive analyses were performed using *compareGroups R* package ^49^.

The final sample size was 1,520 individuals with available information for clinical history, cognitive performance, LTL determinations, and genetic data. Eligible criteria for inclusion in the present study can be found in **Figure 1**.

**Figure 1.**
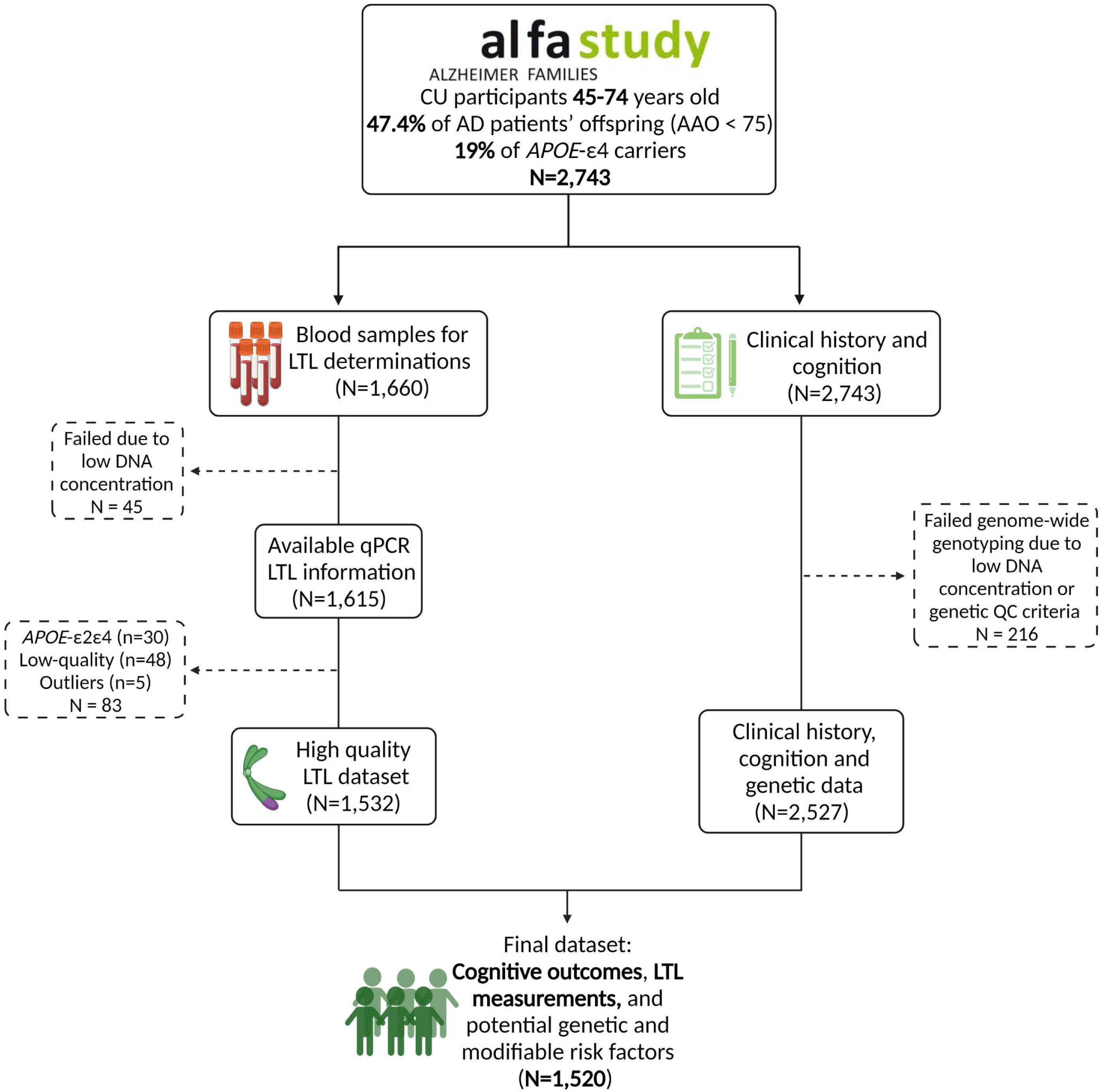
Flow-chart depicting the final sample size of the study. *Legend: AAO, age at symptom onset; AD, Alzheimer’s disease; CU, cognitively unimpaired; LTL, leukocyte telomere length; QC, quality control*.

## 3. Results

### 3.1. Main characteristics of the population by telomere length subgroups

The study sample was classified in two groups of individuals at different stages of biological aging based on the percentile 50^th^ of the rLTL: accelerated biological aging group (i.e., rLTL < percentile 50^th^) and decelerated biological aging group (i.e., rLTL > percentile 50^th^) (see Methods, section 2.6) (**Figure 2**). The accelerated and decelerated aging groups had similar median age at baseline (56 years old) and proportions of women and men. Individuals in the accelerated aging group had higher SBP levels than those in the decelerated aging group (**Table 1**). The distribution of *APOE*-ε4 carriers did not differ between biological aging groups. Individuals in the decelerated aging group had higher polygenic risk scores for AD without *APOE* effect (i.e., PRS-ADno*APOE*). However, individuals with positive parental history of AD were more likely to be classified in the accelerated aging group, and significant differences in the distribution of parental history of AD by parent were observed. Specifically, the percentage of individuals with positive family history of AD in both parents was significantly higher in the accelerated aging group than in the decelerated aging group. Conversely, a higher percentage of positive maternal familial history of AD was observed among individuals in the accelerated aging group. When considering a more stringent parental history encoding (age at symptom onset before 75 years old), no differences were found between the groups, although a similar distribution of parental history of AD by parent was observed (**Table 1**).

**Figure 2.**
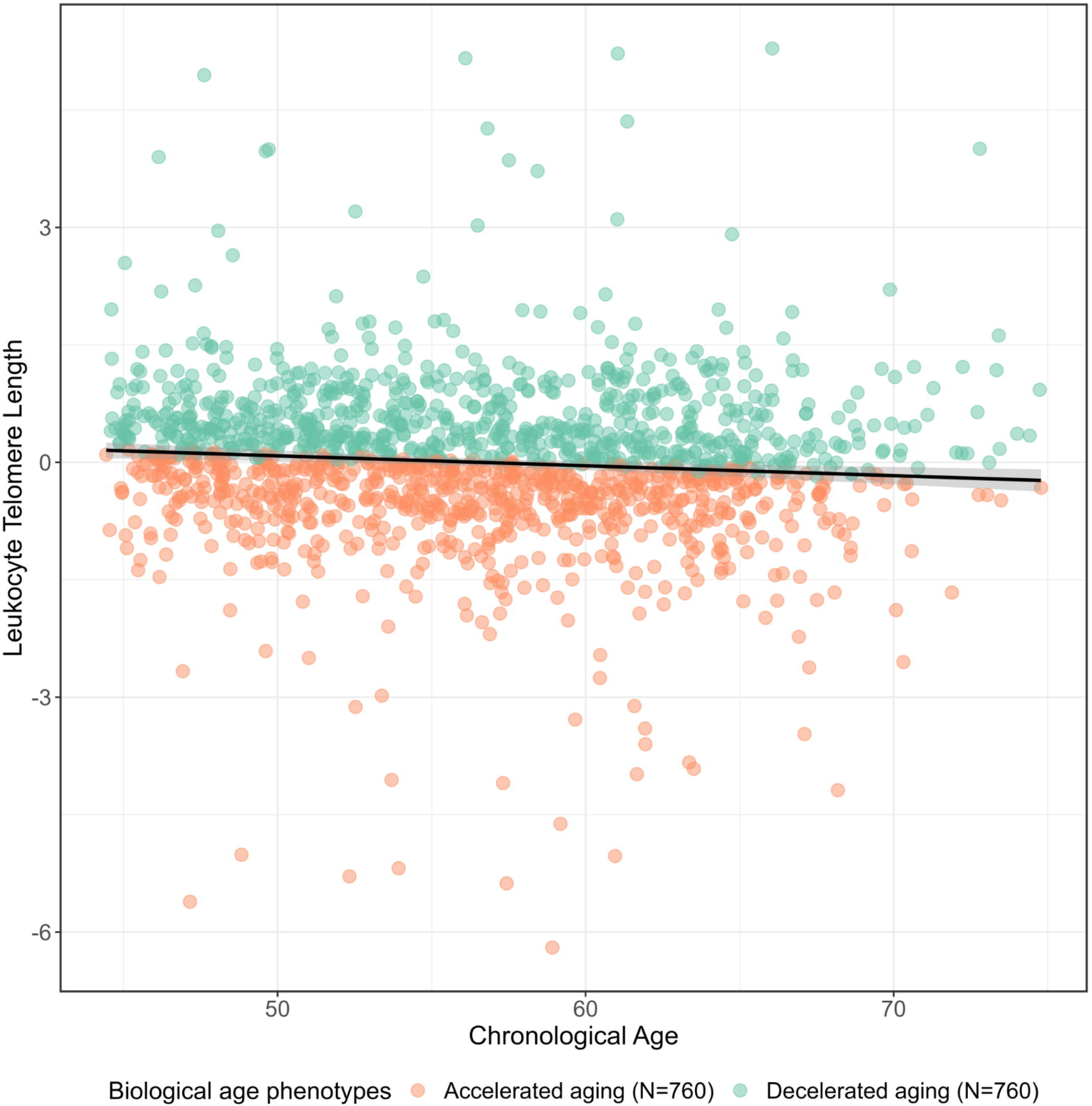
Classification of individuals (N=1,520) in two groups of biological aging based on percentile 50th of residualized leukocyte telomere length.

**Table 1.**
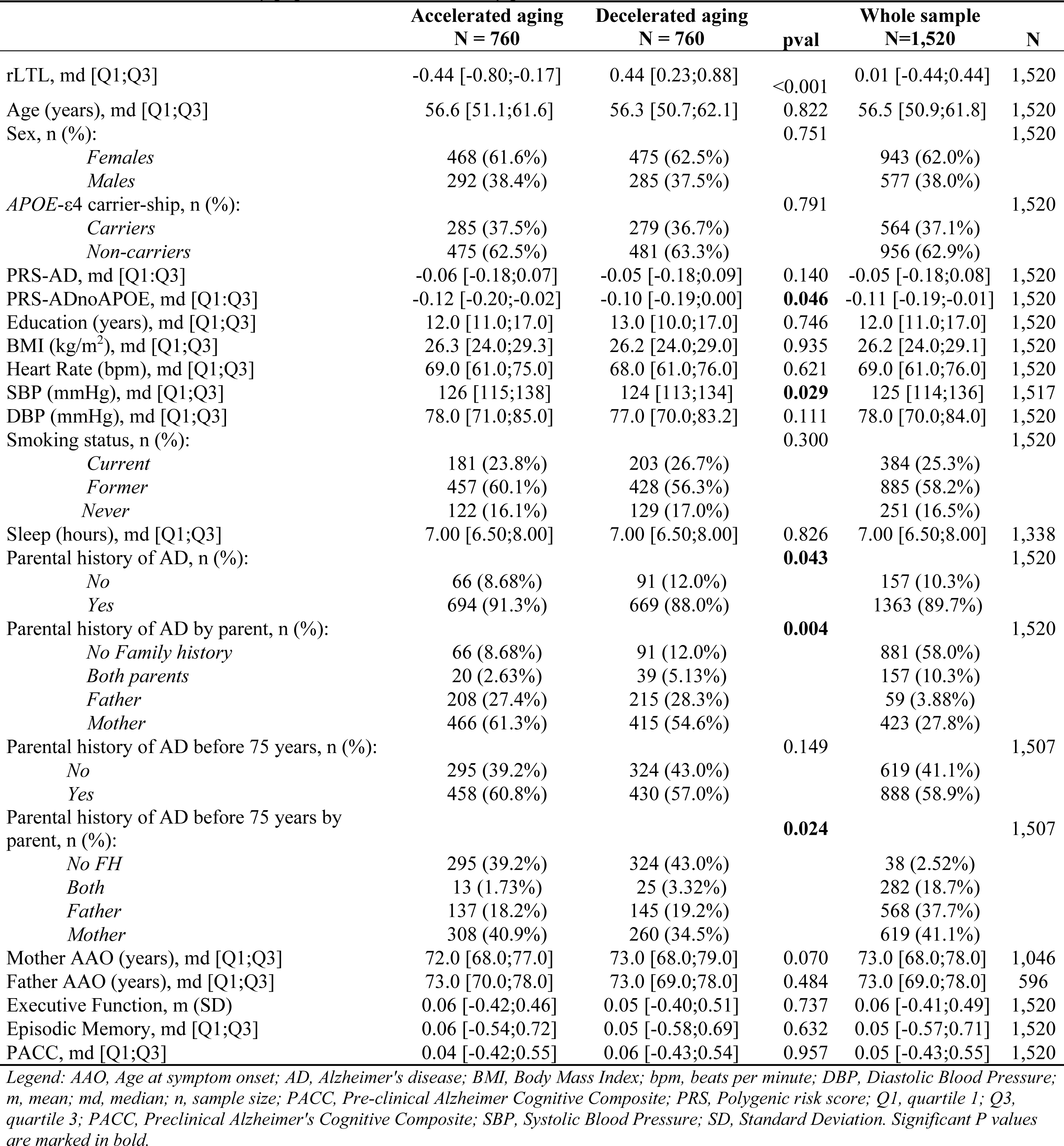
Characteristics of study population at blood draw, by percentile 50^th^ of rLTL.

|  | Accelerated aging<br>N = 760 | Decelerated aging<br>N = 760 | pval | Whole sample<br>N=1,520 | N |
| --- | --- | --- | --- | --- | --- |
| rLTL, md [Q1;Q3] | -0.44 [-0.80;-0.17] | 0.44 [0.23;0.88] | <0.001 | 0.01 [-0.44;0.44] | 1,520 |
| Age (years), md [Q1;Q3] | 56.6 [51.1;61.6] | 56.3 [50.7;62.1] | 0.822 | 56.5 [50.9;61.8] | 1,520 |
| Sex, n (%): |  |  | 0.751 |  | 1,520 |
| *Females* | 468 (61.6%) | 475 (62.5%) |  | 943 (62.0%) |  |
| *Males* | 292 (38.4%) | 285 (37.5%) |  | 577 (38.0%) |  |
| *APOE-ε4* carrier-ship, n (%): |  |  | 0.791 |  | 1,520 |
| *Carriers* | 285 (37.5%) | 279 (36.7%) |  | 564 (37.1%) |  |
| *Non-carriers* | 475 (62.5%) | 481 (63.3%) |  | 956 (62.9%) |  |
| PRS-AD, md [Q1;Q3] | -0.06 [-0.18;0.07] | -0.05 [-0.18;0.09] | 0.140 | -0.05 [-0.18;0.08] | 1,520 |
| PRS-ADnoAPOE, md [Q1;Q3] | -0.12 [-0.20;-0.02] | -0.10 [-0.19;0.00] | **0.046** | -0.11 [-0.19;-0.01] | 1,520 |
| Education (years), md [Q1;Q3] | 12.0 [11.0;17.0] | 13.0 [10.0;17.0] | 0.746 | 12.0 [11.0;17.0] | 1,520 |
| BMI (kg/m<sup>2</sup>), md [Q1;Q3] | 26.3 [24.0;29.3] | 26.2 [24.0;29.0] | 0.935 | 26.2 [24.0;29.1] | 1,520 |
| Heart Rate (bpm), md [Q1;Q3] | 69.0 [61.0;75.0] | 68.0 [61.0;76.0] | 0.621 | 69.0 [61.0;76.0] | 1,520 |
| SBP (mmHg), md [Q1;Q3] | 126 [115;138] | 124 [113;134] | **0.029** | 125 [114;136] | 1,517 |
| DBP (mmHg), md [Q1;Q3] | 78.0 [71.0;85.0] | 77.0 [70.0;83.2] | 0.111 | 78.0 [70.0;84.0] | 1,520 |
| Smoking status, n (%): |  |  | 0.300 |  | 1,520 |
| *Current* | 181 (23.8%) | 203 (26.7%) |  | 384 (25.3%) |  |
| *Former* | 457 (60.1%) | 428 (56.3%) |  | 885 (58.2%) |  |
| *Never* | 122 (16.1%) | 129 (17.0%) |  | 251 (16.5%) |  |
| Sleep (hours), md [Q1;Q3] | 7.00 [6.50;8.00] | 7.00 [6.50;8.00] | 0.826 | 7.00 [6.50;8.00] | 1,338 |
| Parental history of AD, n (%): |  |  | **0.043** |  | 1,520 |
| *No* | 66 (8.68%) | 91 (12.0%) |  | 157 (10.3%) |  |
| *Yes* | 694 (91.3%) | 669 (88.0%) |  | 1363 (89.7%) |  |
| Parental history of AD by parent, n (%): |  |  | **0.004** |  | 1,520 |
| *No Family history* | 66 (8.68%) | 91 (12.0%) |  | 881 (58.0%) |  |
| *Both parents* | 20 (2.63%) | 39 (5.13%) |  | 157 (10.3%) |  |
| *Father* | 208 (27.4%) | 215 (28.3%) |  | 59 (3.88%) |  |
| *Mother* | 466 (61.3%) | 415 (54.6%) |  | 423 (27.8%) |  |
| Parental history of AD before 75 years, n (%): |  |  | 0.149 |  | 1,507 |
| *No* | 295 (39.2%) | 324 (43.0%) |  | 619 (41.1%) |  |
| *Yes* | 458 (60.8%) | 430 (57.0%) |  | 888 (58.9%) |  |
| Parental history of AD before 75 years by parent, n (%): |  |  | **0.024** |  | 1,507 |
| *No FH* | 295 (39.2%) | 324 (43.0%) |  | 38 (2.52%) |  |
| *Both* | 13 (1.73%) | 25 (3.32%) |  | 282 (18.7%) |  |
| *Father* | 137 (18.2%) | 145 (19.2%) |  | 568 (37.7%) |  |
| *Mother* | 308 (40.9%) | 260 (34.5%) |  | 619 (41.1%) |  |
| Mother AAO (years), md [Q1;Q3] | 72.0 [68.0;77.0] | 73.0 [68.0;79.0] | 0.070 | 73.0 [68.0;78.0] | 1,046 |
| Father AAO (years), md [Q1;Q3] | 73.0 [70.0;78.0] | 73.0 [69.0;78.0] | 0.484 | 73.0 [69.0;78.0] | 596 |
| Executive Function, m (SD) | 0.06 [-0.42;0.46] | 0.05 [-0.40;0.51] | 0.737 | 0.06 [-0.41;0.49] | 1,520 |
| Episodic Memory, md [Q1;Q3] | 0.06 [-0.54;0.72] | 0.05 [-0.58;0.69] | 0.632 | 0.05 [-0.57;0.71] | 1,520 |
| PACC, md [Q1;Q3] | 0.04 [-0.42;0.55] | 0.06 [-0.43;0.54] | 0.957 | 0.05 [-0.43;0.55] | 1,520 |

*Legend: AAO, Age at symptom onset; AD, Alzheimer's disease; BMI, Body Mass Index; bpm, beats per minute; DBP, Diastolic Blood Pressure; m, mean; md, median; n, sample size; PACC, Pre-clinical Alzheimer Cognitive Composite; PRS, Polygenic risk score; Q1, quartile 1; Q3, quartile 3; PACC, Preclinical Alzheimer's Cognitive Composite; SBP, Systolic Blood Pressure; SD, Standard Deviation. Significant P values are marked in bold.*

No significant differences were observed among women in the accelerated or decelerated biological aging groups. However, among men, those with paternal family history of AD or both parents diagnosed with the disease were more likely to be classified in the decelerated aging group, while those with positive maternal family history of AD were more prevalent in the accelerated aging group, even when considering a more restrictive parental history encoding (age at symptom onset before 75 years old). Furthermore, men in the accelerated aging group had an earlier maternal symptom onset of AD compared to those in the decelerated aging group. There were no other statistically significant differences observed between the groups. (**Table 2**).

**Table 2.**
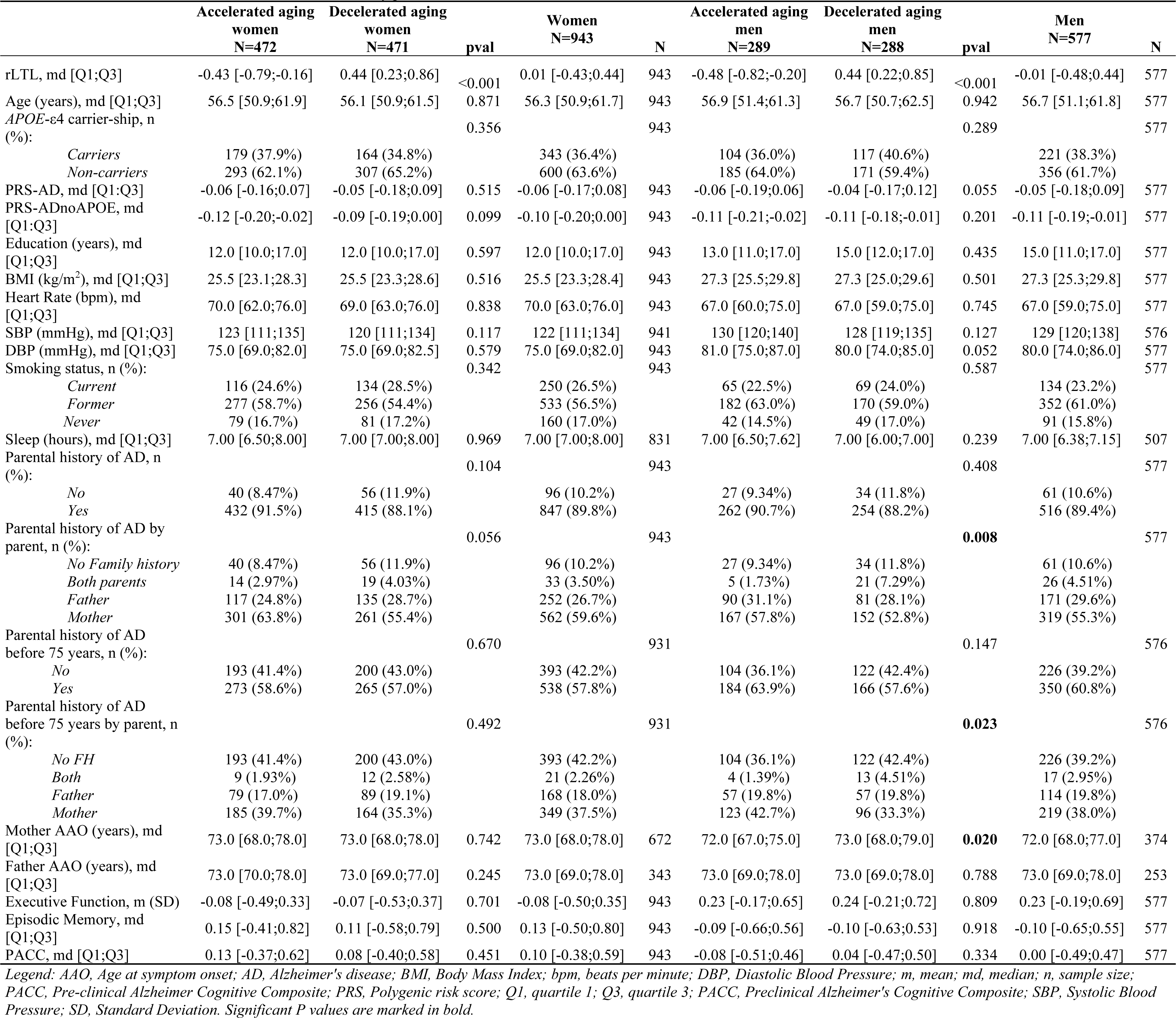
Characteristics of women and men at blood draw, by percentile 50^th^ of rLTL.

### 3.2. Association between rLTL and episodic memory (EM)

The results showed that shorter rLTL at baseline was associated with worse scores on the EM composite, but only among individuals classified in the accelerated aging group [EM composite = β: 0.101, SE: 0.037, FDR-*P* value: .031]. No other statistically significant associations with the EM composite were found in the entire population or among individuals in the decelerated aging group (**Table 3**). This association persisted after adjusting for PRS-ADno*APOE*, SBP and family history of AD (**Supplementary Table 1**). In the analysis with specific scores, shorter rLTL at baseline was associated with worse performance on verbal episodic memory variables related to both immediate and delayed paired and free recall of words from the MBT, but only among individuals within the accelerated aging group. However, after correcting for multiple comparisons only the association with TFR remained significant [TFR = Estimate: 1.039, SE: 1.012, FDR-*P* value: .020] (**Table 3**).

**Table 3.** Association between rLTL and cognitive variables.

|  |  | Whole sample<br>(N=1,520) |  |  |  | Accelerated aging<br>(N=760) |  |  |  | Decelerated aging<br>(N=760) |  |  |  |
| --- | --- | --- | --- | --- | --- | --- | --- | --- | --- | --- | --- | --- | --- |
|  | Outcome | Est.* | SE* | pval | FDR-pval | Est.* | SE* | p-val | FDR-pval | Est.* | SE* | p-val | FDR-pval |
| Memory Binding Test | *TPR*<sup>1</sup> | 1.002 | 1.005 | 0.725 | 0.882 | 1.022 | 1.010 | **0.021** | 0.117 | 0.985 | 1.011 | 0.153 | 0.415 |
|  | *TFR*<sup>1</sup> | 1.000 | 1.006 | 0.991 | 0.991 | 1.039 | 1.012 | **0.001** | **0.020** | 0.979 | 1.013 | 0.117 | 0.415 |
|  | *TDFR*<sup>1</sup> | 1.001 | 1.006 | 0.855 | 0.956 | 1.030 | 1.012 | **0.011** | 0.081 | 0.988 | 1.013 | 0.358 | 0.529 |
|  | *TDPR*<sup>1</sup> | 1.003 | 1.005 | 0.573 | 0.737 | 1.021 | 1.010 | **0.032** | 0.158 | 0.980 | 1.011 | 0.069 | 0.280 |
|  | *SPI*<sup>2</sup> | 0.997 | 1.012 | 0.809 | 0.933 | 1.024 | 1.021 | 0.257 | 0.510 | 0.965 | 1.026 | 0.157 | 0.415 |
|  | **EM composite**<sup>3</sup> | 0.005 | 0.021 | 0.810 | 0.810 | 0.101 | 0.037 | **0.007** | **0.031** | -0.075 | 0.043 | 0.085 | 0.191 |
| WAIS-IV | *Visual Puzzles*<sup>1</sup> | 0.999 | 1.007 | 0.884 | 0.947 | 1.000 | 1.013 | 0.979 | 0.979 | 0.973 | 1.015 | 0.065 | 0.585 |
|  | *Digit Span*<sup>1</sup> | 0.997 | 1.005 | 0.530 | 0.934 | 1.002 | 1.009 | 0.851 | 0.934 | 0.989 | 1.011 | 0.298 | 0.934 |
|  | *Matrix Reasoning*<sup>1</sup> | 0.998 | 1.006 | 0.772 | 0.934 | 0.992 | 1.011 | 0.499 | 0.934 | 0.993 | 1.013 | 0.610 | 0.934 |
|  | *Similarities*<sup>1</sup> | 1.004 | 1.006 | 0.525 | 0.934 | 1.009 | 1.010 | 0.382 | 0.934 | 0.991 | 1.011 | 0.435 | 0.934 |
|  | *Coding*<sup>1</sup> | 0.990 | 1.003 | **0.002** | 0.073 | 0.998 | 1.006 | 0.726 | 0.934 | 0.984 | 1.007 | **0.012** | 0.140 |
|  | **EF composite**<sup>3</sup> | -0.010 | 0.014 | 0.439 | 0.801 | -0.004 | 0.024 | 0.858 | 0.914 | -0.044 | 0.026 | 0.094 | 0.608 |
| Semantic Fluency | *Animals in one minute*<sup>1</sup> | 1.005 | 1.006 | 0.389 | 0.772 | 1.002 | 1.010 | 0.864 | 0.864 | 1.003 | 1.011 | 0.789 | 0.864 |
| **Preclinical Alzheimer Cognitive Composite**<sup>3</sup> |  | -0.002 | 0.016 | 0.890 | 0.991 | 0.053 | 0.028 | 0.061 | 0.275 | -0.046 | 0.033 | 0.164 | 0.491 |

Legend: EM, episodic memory; EF, executive functioning; FDR: False Discovery Rate; TPR, Total paired recall; TFR, Total free recall; TDFR, Total delayed free recall; TDPR, Total delayed paired recall; SE, Standard Error; SPI, Semantic Proactive interference; WAIS-IV, Wechsler Adult Intelligence Scale- Fourth Edition.
Significant P values are marked in bold. All continuous predictors are mean-centered and scaled by 1 standard deviation.
(1) Outcomes modeled under Poisson distribution.
(2) Outcomes modeled under Quasibinomial distribution.
(3) Outcomes modeled under Normal distribution. Models were adjusted by age, sex and APOE-ε4 status.
(\*) Raw effects (estimates and SE) were exponentiated for outcomes modeled under Poisson and Quasibinomial distributions.

In the entire sample, a suggestive rLTL-sex interaction effect was found on EM [*P* value-interaction: .065] (**Supplementary Figure 4**, **Supplementary Table 2**). Similarly, a statistically significant sex-interaction effect was observed among individuals in the accelerated aging group [*P* value-interaction: .043] (**Figure 3**, **Supplementary Table 2**). Specifically, shorter rLTL at baseline had a detrimental effect on the EM composite among women in the accelerated aging group [EM composite = β: 0.151, SE: 0.045, *P* value: .007], whereas longer rLTL had a suggestive detrimental effect among men in the decelerated aging group [EM composite = β: −0.150, SE: 0.069, *P* value: .030] (**Table 4**).

**Figure 3.**
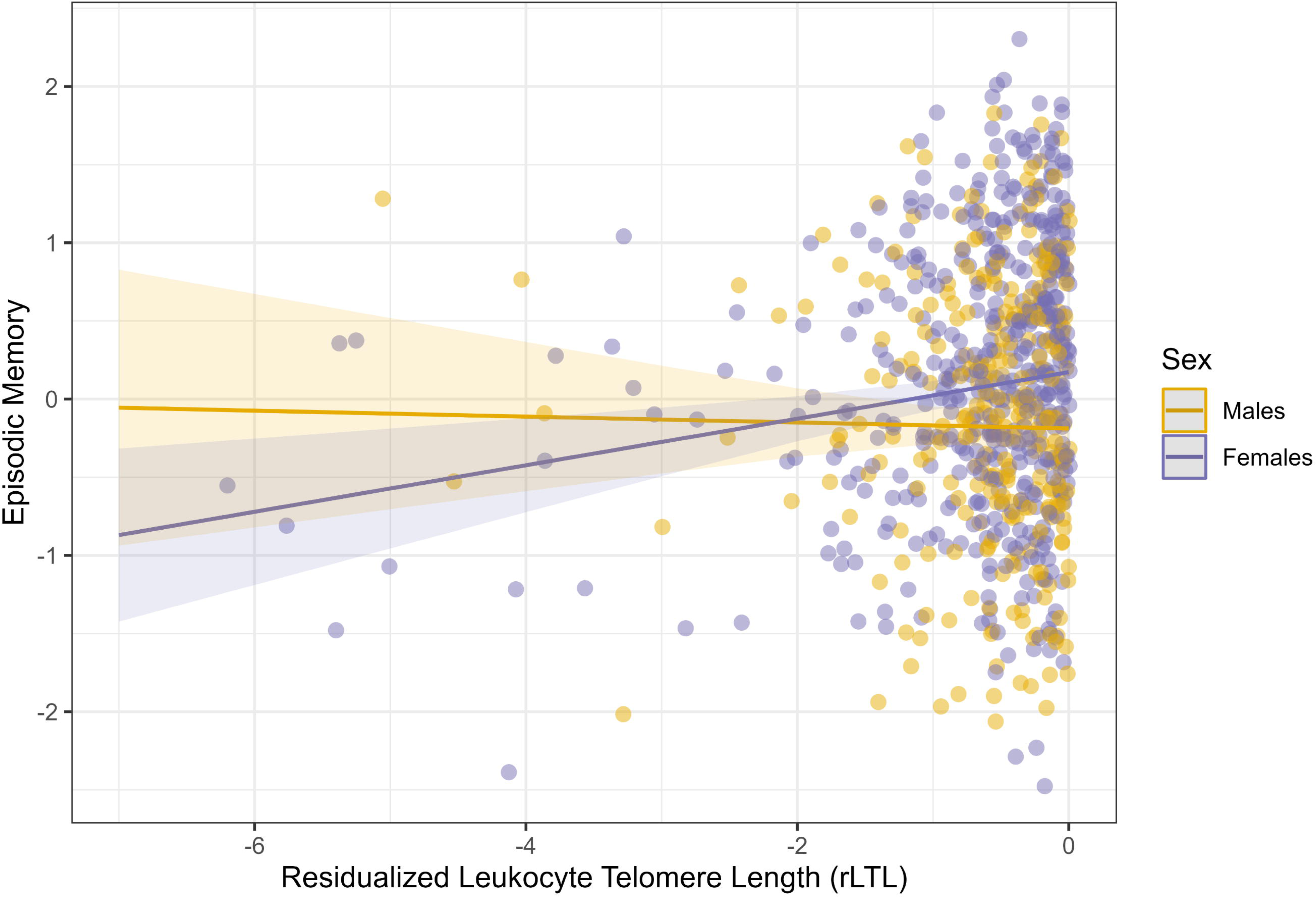
Significant interaction effect of sex and residualized leukocyte telomere length on episodic memory among individuals at accelerated aging (N=760). *[P-Interaction = 0.043]*

**Table 4.** Association between rLTL and cognitive variables stratifying by sex.

|  |  | Women<br>(N=943) |  |  |  | Women at accelerated aging<br>(N=472) |  |  |  | Women at decelerated aging<br>(N=471) |  |  |  | Men<br>(N=577) |  |  |  | Men at accelerated aging<br>(N=289) |  |  |  | Men at decelerated aging<br>(N=288) |  |  |  |
| --- | --- | --- | --- | --- | --- | --- | --- | --- | --- | --- | --- | --- | --- | --- | --- | --- | --- | --- | --- | --- | --- | --- | --- | --- | --- |
|  | Outcome |  |  |  |  |  |  |  |  |  |  |  |  |  |  |  |  |  |  |  |  |  |  |  |  |
|  |  | Est.* | SE* | p-val | FDR-pval | Est.* | SE* | p-val | FDR-pval | Est.* | SE* | p-val | FDR-pval | Est.* | SE* | p-val | FDR-pval | Est.* | SE* | p-val | FDR-pval | Est.* | SE* | p-val | FDR-pval |
| Memory Binding Test | TPR<sup>1</sup> | 1.007 | 1.007 | 0.264 | 0.510 | 1.032 | 1.012 | **0.008** | 0.081 | 0.995 | 1.014 | 0.712 | 0.882 | 0.991 | 1.009 | 0.295 | 0.510 | 1.001 | 1.017 | 0.967 | 0.991 | 0.964 | 1.018 | **0.045** | 0.201 |
|  | TFR<sup>1</sup> | 1.005 | 1.008 | 0.539 | 0.714 | 1.057 | 1.015 | **0.000** | **0.008** | 0.985 | 1.017 | 0.365 | 0.529 | 0.988 | 1.011 | 0.274 | 0.510 | 0.999 | 1.020 | 0.977 | 0.991 | 0.967 | 1.023 | 0.130 | 0.415 |
|  | TDFR<sup>1</sup> | 1.009 | 1.008 | 0.267 | 0.510 | 1.051 | 1.015 | **0.001** | **0.013** | 0.996 | 1.016 | 0.803 | 0.933 | 0.984 | 1.011 | 0.138 | 0.415 | 0.982 | 1.020 | 0.347 | 0.529 | 0.968 | 1.022 | 0.143 | 0.415 |
|  | TDPR<sup>1</sup> | 1.008 | 1.007 | 0.204 | 0.459 | 1.031 | 1.012 | **0.010** | 0.081 | 0.991 | 1.014 | 0.523 | 0.713 | 0.992 | 1.009 | 0.346 | 0.529 | 0.997 | 1.017 | 0.871 | 0.956 | 0.958 | 1.019 | **0.020** | 0.117 |
|  | SPI<sup>2</sup> | 1.010 | 1.015 | 0.517 | 0.713 | 1.035 | 1.025 | 0.166 | 0.416 | 0.969 | 1.033 | 0.333 | 0.529 | 0.968 | 1.025 | 0.193 | 0.457 | 1.002 | 1.046 | 0.971 | 0.991 | 0.948 | 1.053 | 0.293 | 0.510 |
|  | EM composite<sup>3</sup> | 0.033 | 0.027 | 0.228 | 0.342 | 0.151 | 0.045 | **0.001** | **0.007** | -0.039 | 0.057 | 0.495 | 0.636 | -0.050 | 0.035 | 0.154 | 0.278 | -0.022 | 0.067 | 0.747 | 0.810 | -0.150 | 0.069 | **0.030** | 0.090 |
| WAIS-IV<br>Test | Visual Puzzles<sup>1</sup> | 1.002 | 1.009 | 0.826 | 0.934 | 1.003 | 1.016 | 0.839 | 0.934 | 0.978 | 1.019 | 0.251 | 0.934 | 0.994 | 1.011 | 0.617 | 0.934 | 0.994 | 1.021 | 0.787 | 0.934 | 0.965 | 1.023 | 0.118 | 0.661 |
|  | Digit Span<sup>1</sup> | 1.002 | 1.007 | 0.710 | 0.934 | 1.005 | 1.012 | 0.640 | 0.934 | 0.993 | 1.014 | 0.613 | 0.934 | 0.986 | 1.008 | 0.095 | 0.661 | 0.990 | 1.016 | 0.524 | 0.934 | 0.977 | 1.017 | 0.174 | 0.871 |
|  | Matrix Reasoning<sup>1</sup> | 0.994 | 1.008 | 0.490 | 0.934 | 0.990 | 1.014 | 0.484 | 0.934 | 0.988 | 1.017 | 0.487 | 0.934 | 1.005 | 1.010 | 0.645 | 0.934 | 0.995 | 1.019 | 0.813 | 0.934 | 1.001 | 1.021 | 0.971 | 0.979 |
|  | Similarities<sup>1</sup> | 1.000 | 1.007 | 0.946 | 0.979 | 1.005 | 1.012 | 0.667 | 0.934 | 0.989 | 1.014 | 0.442 | 0.934 | 1.008 | 1.009 | 0.349 | 0.934 | 1.015 | 1.017 | 0.386 | 0.934 | 0.992 | 1.018 | 0.671 | 0.934 |
|  | Coding<sup>1</sup> | 0.989 | 1.004 | **0.009** | 0.135 | 0.998 | 1.007 | 0.764 | 0.934 | 0.976 | 1.008 | **0.004** | 0.079 | 0.992 | 1.005 | 0.111 | 0.661 | 0.996 | 1.010 | 0.665 | 0.934 | 0.997 | 1.011 | 0.777 | 0.934 |
|  | EF composite<sup>3</sup> | -0.007 | 0.017 | 0.695 | 0.914 | -0.003 | 0.030 | 0.914 | 0.914 | -0.049 | 0.033 | 0.135 | 0.608 | -0.018 | 0.023 | 0.445 | 0.801 | -0.013 | 0.044 | 0.771 | 0.914 | -0.044 | 0.045 | 0.330 | 0.801 |
| Semantic Fluency | Animals in one minute<sup>1</sup> | 0.995 | 1.007 | 0.476 | 0.772 | 0.994 | 1.012 | 0.600 | 0.772 | 0.991 | 1.014 | 0.544 | 0.772 | 1.023 | 1.009 | **0.012** | 0.111 | 1.016 | 1.017 | 0.354 | 0.772 | 1.017 | 1.018 | 0.332 | 0.772 |
| Preclinical Alzheimer Cognitive Composite<sup>3</sup> |  | ~0 | 0.020 | 0.987 | 0.991 | 0.071 | 0.034 | **0.038** | 0.275 | -0.048 | 0.043 | 0.257 | 0.530 | -0.008 | 0.027 | 0.757 | 0.991 | -0.001 | 0.051 | 0.991 | 0.991 | -0.054 | 0.052 | 0.295 | 0.530 |

Legend: EM, episodic memory; EF, executive functioning; FDR: False Discovery Rate; TPR, Total paired recall; TFR, Total free recall; TDFR, Total delayed free recall; TDPR, Total delayed paired recall; SE, Standard Error; SPI, Semantic Proactive interference; WAIS-IV, Wechsler Adult Intelligence Scale- Fourth Edition.
Significant P values are marked in bold. All continuous predictors are mean-centered and scaled by 1 standard deviation.
(1) Outcomes modeled under Poisson distribution.
(2) Outcomes modeled under Quasibinomial distribution.
(3) Outcomes modeled under Normal distribution. Models were adjusted by age, sex and APOE-ε4 status.
(\*) Raw effects (estimates and SE) were exponentiated for outcomes modeled under Poisson and Quasibinomial distributions.

Regarding specific cognitive variables, shorter rLTL at baseline had significant detrimental effects on both TFR and TDFR among women included in the accelerated aging group [TFR = Estimate: 1.057, SE: 1.015, FDR-*P* value: .008; TDFR = Estimate: 1.051, SE: 015, FDR-*P* value: .013]. However, nominal statistically significant associations between longer rLTL and lower performance in TPR and TDPR were observed among men in the decelerated aging group [TPR= Estimate: 0.991, SE: 1.018, *P* value: .045; TDPR= Estimate: 0.948, SE: 1.053, *P* value: .020] (**Table 4**).

No other significant interaction effects on the EM composite were observed between rLTL and the genetic risk of AD, including *APOE*-ε4 status,PRS-ADno*APOE,* PRS-CSFAβ or PRS-CSFpTau (**Supplementary Tables 3-6**).

### 3.3. Association between rLTL, executive function and semantic fluency

In our population, no associations were found between rLTL and the EF composite in the entire sample, nor in subgroup analyses performed independently in the biological aging subgroups (**Table 3**). These associations were not changed after fully adjusting the models (**Supplementary Table 1**). Additionally, on the EF composite, rLTL had no interactive effect with sex, *APOE*-ε4, PRS-ADno*APOE*, PRS-CSFAβ or PRS-CSFpTau (**Supplementary Tables 2-6**).

Regarding specific subtests on the EF domain, nominally statistically significant associations were found between longer rLTL and worse performance on WAIS-IV Coding, both in the whole sample [Coding = Estimate: 0.990, SE: 1.003, *P* value: .002] and among individuals in the decelerated aging group [Coding = Estimate: 0.984, SE: 1.007, *P* value: .012] (**Table 3**). These associations were mainly driven by women (**Table 4**).

Additionally, a nominally significant association was observed among men, where longer rLTL was associated with better semantic fluency [Semantic Fluency = Estimate: 1.023, SE: 1.009, *P* value: .012] (**Table 4**).

### 3.4. Association between rLTL and pre-clinical Alzheimer cognitive composite

Shorter rLTL at baseline was nominally associated with lower scores in the PACC, but only among women in the accelerated aging group [PACC = β: 0.071, SE: 0.034, *P* value: .038] (**Table 4**). In post-hoc sensitivity analyses, adjustment by further potential confounders (i.e., PRS-ADno*APOE*, SBP and family history of AD) revealed a borderline nominally statistically significant association between shorter rLTL and lower scores in the PACC among individuals in the accelerated aging group [PACC = β: 0.044, SE: .022, *P* value: .049] (**Supplementary Table 1**). No other statistically significant associations were found (**Table 3-4**).

## 4. Discussion

The present study explored the cross-sectional association between rLTL and cognitive performance among 1,520 cognitively healthy individuals from the ALFA study. We found that shorter rLTL at baseline was associated with worse EM only among individuals with shorter telomeres than expected for their age (i.e., accelerated aging group [N = 760]). A sex-interaction effect on the EM composite was found among individuals at accelerated aging: shorter rLTL at baseline was associated with worse performance on the EM domain only among women classified in the accelerated aging group (N = 472), whereas no significant association was found among men in the accelerated aging group (N = 289). Although no sex-interactive effect was found among individuals at decelerated aging, longer rLTL had a detrimental effect on the EM composite among men (N = 288) but not women at decelerated aging (N = 471). To our knowledge, this is the first study examining sex-differences in LTL-cognition associations.

Regarding cognitive variables, shorter rLTL at baseline had significant detrimental effects on both TFR and TDFR among women included in the accelerated aging group. However, among men in the decelerated aging group, associations between longer rLTL and lower performance in TPR and TDPR were observed. Remarkably, only free recall measurements remained significant after correction for multiple comparisons. Cued recall measurements are considered better predictors of memory impairment due to AD dementia since they remain intact during normal aging ^50^, being able to distinguish people living with AD from age-matched controls ^51–54^. Nevertheless, free recall measurements have been found to be more sensitive for prediction of AD dementia specifically during pre-clinical stages ^55–57^. Interestingly, the delayed free recall of the second list of the MBT has been associated with amyloid burden in precuneus in clinical normal elderly ^58^. Moreover, worse performance in TDFR but not cued recall was found in prodromal amyloid positive individuals without degeneration, while both TDFR and TDPR worsened in more advanced biomarker stages with amyloid burden and neurodegeneration ^59^. In accordance with these findings, it has been described a sequence of memory impairment in the course of AD in which the first change occurs in free recall, while additional impairment in cued recall (i.e. paired recall trials in the MBT) occur later on ^60, 61^. Additionally, free recall is more affected by aging than cued recall or recognition ^62^, since it relies on active self-guided processes that are dependent on the function of the frontal lobe ^63^, which is known to be the brain region primarily affected by aging ^64, 65^. Thus, our observed associations between free recall measures and rLTL in CU individuals at increased risk of AD may relate to an exacerbated cognitive aging and/or the effect of early AD pathological changes in some individuals ^66^.

Regarding the EF domain, the association between longer rLTL and poorer processing speed (specifically, WAIS-IV Coding) found to be nominally statistically significant, in both the entire sample and the decelerated aging group. However, given the lack of other significant associations related to executive performance in our sample, this finding should be interpreted with caution. In contrast, longer rLTL was nominally associated with better semantic fluency, but only among men (N = 577). Nominal significant associations were also found between shorter rLTL and worse PACC among women in the accelerated aging group. However, since PACC was computed by averaging scores coming from variables of MBT, WAIS-IV, and semantic fluency, we suspect this result was mainly driven by results in the EM domain and countered by the opposite associations encountered on the EF domain.

Previous studies have reported associations between longer LTL and better performance in a variety of cognitive domains and general cognition in meta-analytic settings ^24, 67^. Moreover, shorter LTL predicted 20-year memory decline among 880 dementia-free participants from a population-based study ^68^. In the UKBiobank, longer LTL was significantly associated with faster reaction time, higher fluid intelligence and higher numeric memory ^22^. In contrast, longer telomeres at baseline had been associated with faster EF decline among AD biomarker-positive individuals and carriers of the *APOE*-ε4 allele ^27^, whereas longer LTL predicted worse episodic memory among *APOE*-ε4 carriers ^69^. Thus, whether longer telomeres play a protective or perjudicial role on cognitive resilience remains unclear, specially in the presence of other risk factors for AD. Interestingly, previous observational studies have described non-linear associations between LTL and AD risk ^18, 19^, with different directions of associations among *APOE*-ε4 carriers and non-carriers ^18^. However, no effect modification by *APOE*-ε4 status or the polygenic liability for AD were observed in the associations between rLTL and the outcomes of the present study. Together, these results suggest complex heterogeneity in LTL mechanisms that may appear to change over the AD spectrum and the *continuum* of the disease.

Given the aforementioned results, biological aging due to telomere shortening could interact with the AD pathophysiological process and contribute to cognitive vulnerability since the earliest pre-clinical stages of the disease ^22, 70, 71^. Importantly, only critically short telomeres are able to trigger cellular senescence ^72^ and senescence phenotypes resulting from telomere shortening have been detected in microglial cells from post-mortem samples of people living with AD, indicating a possible link between telomere shortening and AD pathogenesis ^73^. Additionally, telomere shortening has been associated with reduced hippocampal progenitor cell proliferation and changes in gene expression that affect cognitive function ^74^. Conversely, a recent Mendelian Randomization study conducted in our sample showed genetically predicted longer LTL was associated with lower levels of cerebrospinal fluid (CSF) Aβ and higher levels of CSF NfL only among *APOE*-ɛ4 non-carriers. On the contrary, inheriting longer LTL was associated with lower levels of CSF p-tau among individuals with high polygenic risk scores of AD ^75^. The complex and dynamic relationship between biological aging and AD pathology through the disease *continuum* may partially explain the non-linear associations between LTL, cognitive performance, and AD risk, as suggested by the conflicting associations observed.

Our study is not without limitations. Our cohort is rather selected and composed of middle-aged CU individuals. Therefore, individuals at high risk of AD might have already shown signs of cognitive decline at recruitment, which was an exclusion criterion of the present study. Furthermore, participants in the ALFA study showed a low prevalence of other common comorbidities ^28^ and therefore our results might not be generalizable to the general population. On the other hand, sex-interactions, and stratified analyses by sex drawback from our sex-unbalanced sample (i.e., 62% women vs. 38% males), which could explain lack of significant sex-interactions in the entire sample as well as associations among men. Importantly, given its cross-sectional design, our results on cognitive performance might reflect a transient stage within the AD pathological process. Thus, longitudinal studies with greater sample sizes will be warranted to unravel the effect of telomere maintenance on cognitive performance during mid-life. Finally, those associations that did not survive FDR multiple comparisons correction should be interpreted with caution.

However, this study also exhibits multiple strengths. Our study is based on a robust and well-characterized cohort of CU middle-aged individuals. This included the extensive characterization of cognitive outcomes in the ALFA participants compared to the general population. As far as we know, no prior studies have explored sex-specific patterns of telomere effects on cognitive performance in a similar cohort of CU individuals at increased risk of developing AD. This implies our findings should be replicated in larger cohorts including people living with AD at different stages of the disease. Similarly, the follow-up of ALFA participants and further prospective observational studies are required to better understand such observations.

In conclusion, this cross-sectional study shows a non-linear association between LTL and cognitive performance, with heterogeneous associations between telomere length and different cognitive domains. Shorter telomeres at baseline were associated with poorer memory performance among individuals at accelerated biological aging. Moreover, our findings indicate sex-specific effects, with women at accelerated aging experiencing more pronounced cognitive impairment than their male counterparts. Our results suggest sex-specific cognitive vulnerability associated with telomere homeostasis in our cohort.

## Data Availability

All data produced in the present study are available upon reasonable request to the authors.

## List of supplementary figures

**Supplementary Figure 1.** The non-linear association between residualized leukocyte telomere length (rLTL) and Episodic Memory composite was investigated by including (a) one-knot, (b) two-knots, and (c) three-knots natural splines. The best-fitted model was selected using the Bayesian Information Criteria (BIC). Lower BIC values represent better fitted-models.

**Supplementary Figure 2.** The non-linear association between residualized leukocyte telomere length (rLTL) and Executive Function composite was investigated by including (a) one-knot, (b) two-knots, and (c) three-knots natural splines. The best-fitted model was selected using the Bayesian Information Criteria (BIC). Lower BIC values represent better fitted-models.

**Supplementary Figure 3.** The non-linear association between residualized leukocyte telomere length (rLTL) and Preclinical Alzheimer Cognitive composite was investigated by including (a) one-knot, (b) two-knots, and (c) three-knots natural splines. The best-fitted model was selected using the Bayesian Information Criteria (BIC). Lower BIC values represent better fitted-models.

**Supplementary Figure 4.** Interaction effect between sex and rLTL on episodic memory in the entire sample. *[P-Interaction = 0.06]*

## List of supplementary tables

**Supplementary Table 1.** Fully adjusted models examining the association between rLTL and the main cognitive outcomes of the study. *Legend: AD, Alzheimer’s disease; PRS, Polygenic risk score; rLTL, residualized leukocyte telomere length; SE, standard error. Significant P values are marked in bold. All continuous predictors are mean-centered and scaled by 1 standard deviation*.

**Supplementary Table 2.** Interactive effect of rLTL and sex on the main outcomes of the study. *Legend: AD, Alzheimer’s disease; rLTL, residualized leukocyte telomere length; SE, standard error. Significant P values are marked in bold. All continuous predictors are mean-centered and scaled by 1 standard deviation*.

**Supplementary Table 3.** Interactive effect of rLTL and *APOE*-ε4 carriership on the main outcomes of the study. *Legend: AD, Alzheimer’s disease; rLTL, residualized leukocyte telomere length; SE, standard error. Significant P values are marked in bold. All continuous predictors are mean-centered and scaled by 1 standard deviation*.

**Supplementary Table 4.** Interactive effect of rLTL and PRS-ADnoAPOE on the main outcomes of the study. *Legend: AD, Alzheimer’s disease; PRS, Polygenic risk score; rLTL, residualized leukocyte telomere length; SE, standard error. Significant P values are marked in bold. All continuous predictors are mean-centered and scaled by 1 standard deviation*.

**Supplementary Table 5.** Interactive effect of rLTL and PRS-CSFAβ on the main outcomes of the study. *Legend: Aβ, amyloid-β; CSF, cerebrospinal fluid; PRS, polygenic risk score; rLTL, residualized leukocyte telomere length; SE, standard error. Significant P values are marked in bold. All continuous predictors are mean-centered and scaled by 1 standard deviation*.

**Supplementary Table 6.** Interactive effect of rLTL and PRS-CSFpTau on the main outcomes of the study. *Legend: CSF, cerebrospinal fluid; PRS, polygenic risk score; pTau, phosphorylated tau; rLTL, residualized leukocyte telomere length; SE, standard error. Significant P values are marked in bold. All continuous predictors are mean-centered and scaled by 1 standard deviation*.

## Acknowledgements

This publication is part of the ALFA study. The authors would like to express their most sincere gratitude to the ALFA project participants and relatives without whom this research would have not been possible. Collaborators of the ALFA study are: Müge Akinci, Federica Anastasi, Annabella Beteta, Raffaele Cacciaglia, Lidia Canals, Alba Cañas, Carme Deulofeu, Maria Emilio, Irene Cumplido-Mayoral, Marta del Campo, Carme Deulofeu, Ruth Dominguez, Maria Emilio, Sherezade Fuentes, Marina García, Armand González-Escalante, Laura Hernández, Gema Huesa, Jordi Huguet, Laura Iglesias, Esther Jiménez, David López-Martos, Paula Marne, Tania Menchón, Paula Ortiz-Romero, Eleni Palpatzis, Wiesje Pelkmans, Albina Polo, Sandra Pradas, Iman Sadeghi, Mahnaz Shekari, Lluís Solsona, Anna Soteras, Laura Stankeviciute, Núria Tort-Colet and Marc Vilanova. We would like to additionally express our sincere gratitude to Prof. Immaculata de Vivo for her invaluable assistance in acquiring the telomere length data used in this study.

## Consent statement

The study was conducted in accordance with the directives of the Spanish Law 14/2007, of 3rd of July, on Biomedical Research (Ley 14/2007 de Investigación Biomédica). The ALFA study protocol was approved by the Independent Ethics Committee Parc de Salut Mar Barcelona and registered at Clinicaltrials.gov (Identifier: NCT01835717). All participants accepted the study procedures by signing the study’s informed consent form that had also been approved by the same IRB.

