## Supplementary figures and images for "Association between telomere length and cognitive function among cognitively unimpaired individuals at risk of Alzheimer’s disease"

### Supplementary Figure 1

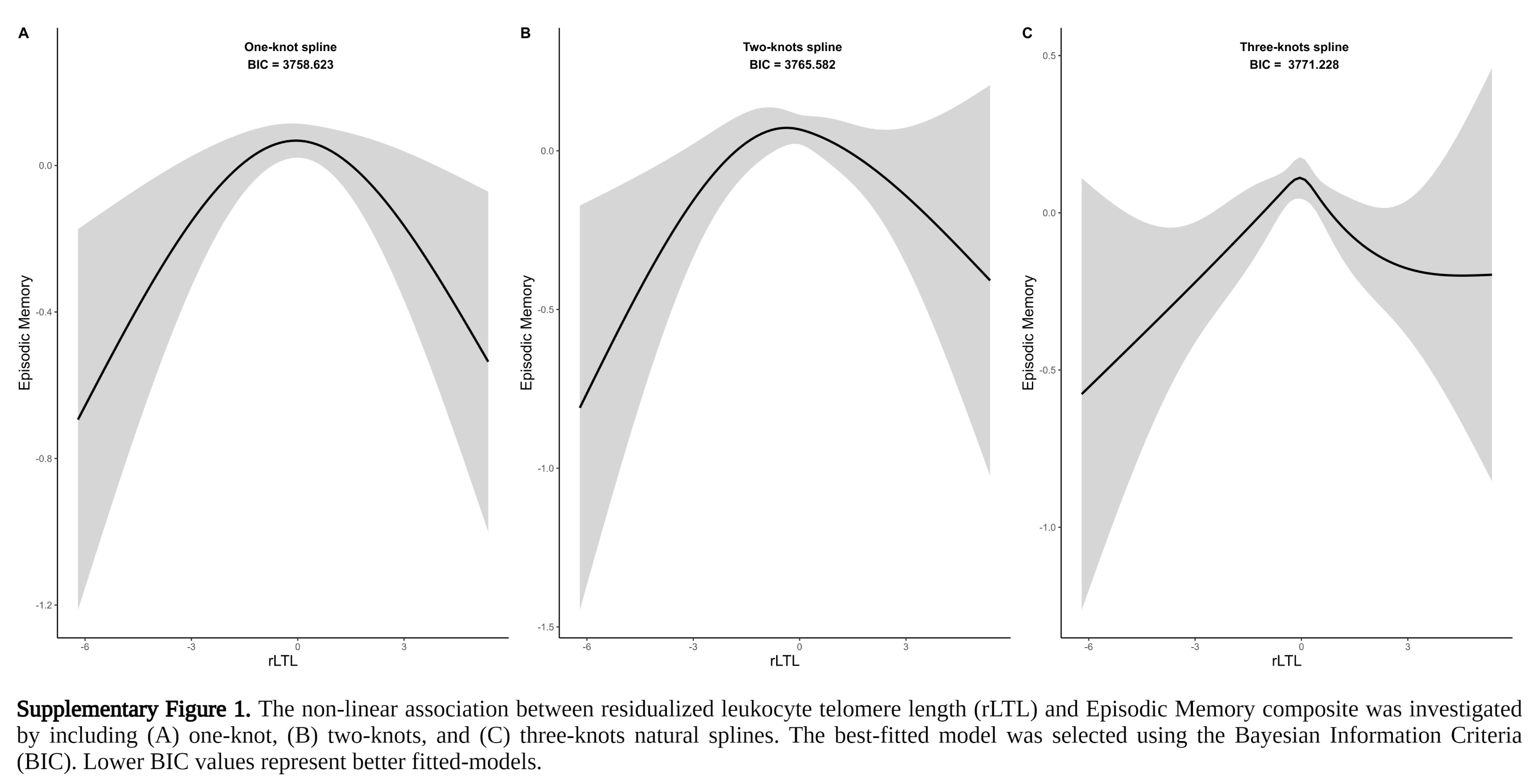

### Supplementary Figure 2

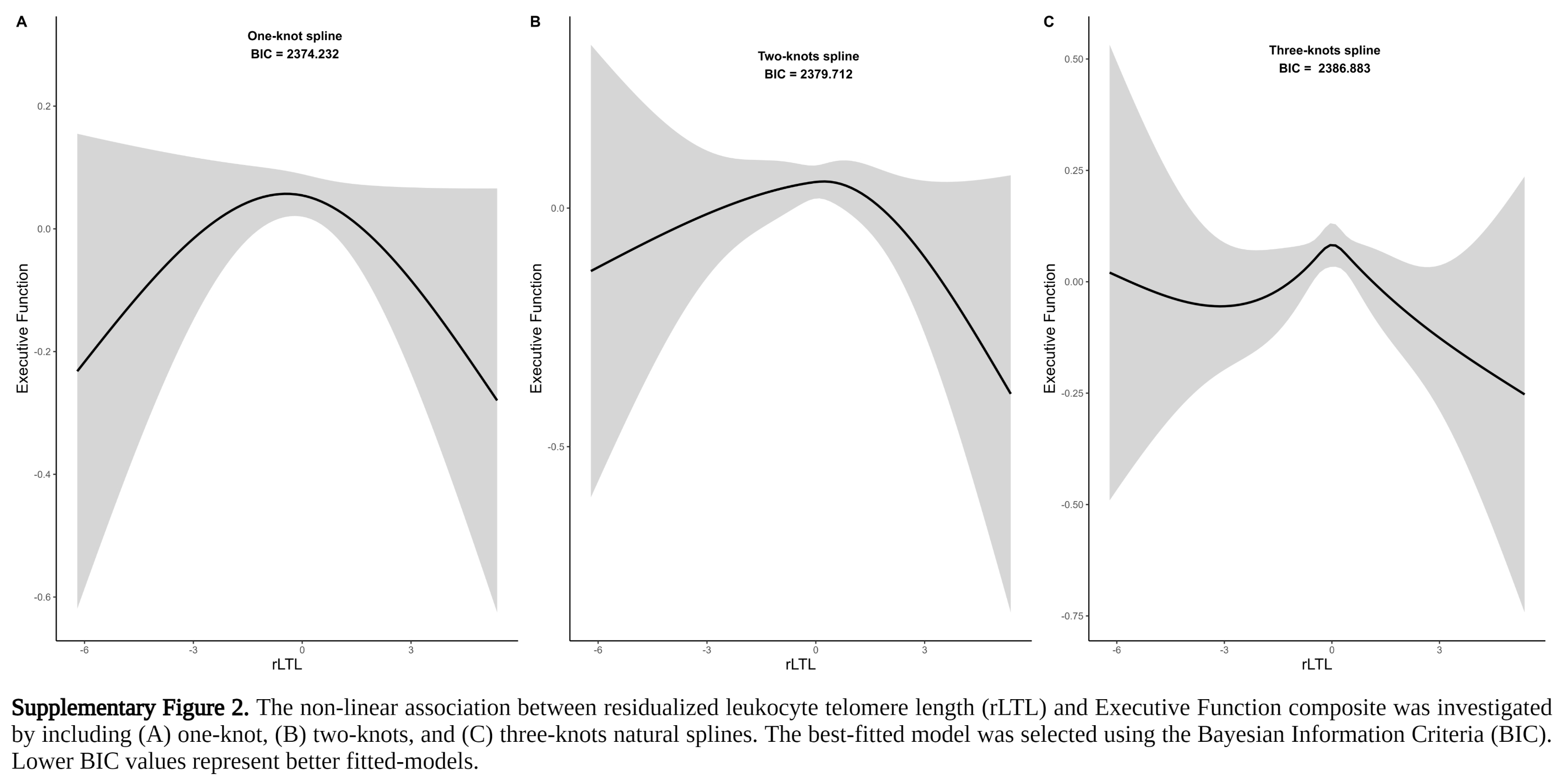

### Supplementary Figure 3

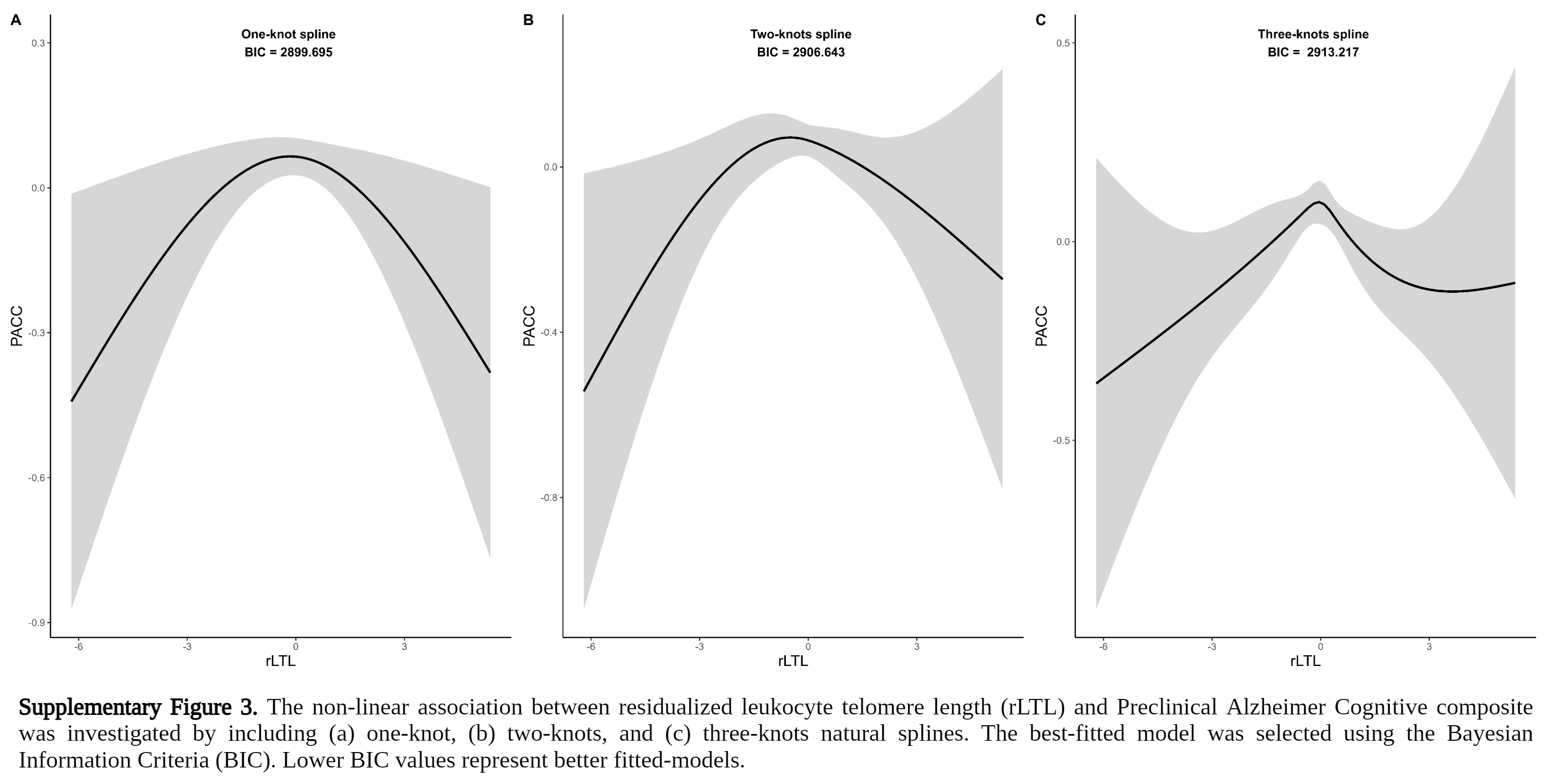

### Supplementary Figure 4

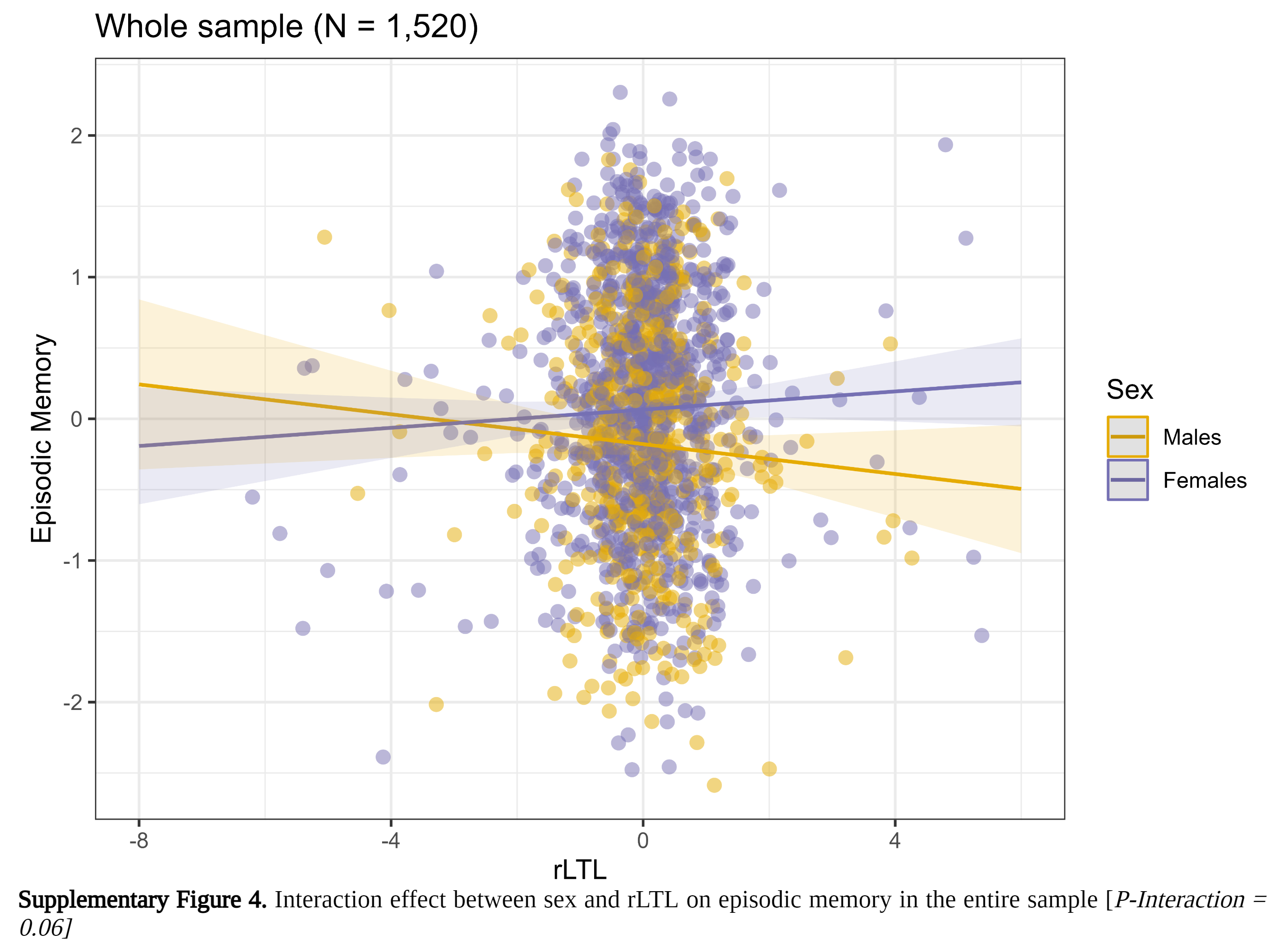
